# Toxic Metals Impact Gut Microbiota and Metabolic Risk in Five African-Origin Populations

**DOI:** 10.1101/2024.10.07.24315016

**Authors:** Julianne A. Jorgensen, Candice Choo-Kang, Luyu Wang, Lina Issa, Jack A. Gilbert, Gertrude Ecklu-Mensah, Amy Luke, Kweku Bedu-Addo, Terrence Forrester, Pascal Bovet, Estelle V. Lambert, Dale Rae, Maria Argos, Tanika N. Kelly, Robert M. Sargis, Lara R. Dugas, Yang Dai, Brian T. Layden

## Abstract

Exposure to toxic metals impacts obesity and type 2 diabetes (T2DM) risk. Yet, the underlying mechanisms remain largely unknown. Gut microbiota has been strongly associated with progression of cardiometabolic risk. To determine whether high metal exposures and gut dysbiosis interact to promote metabolic dysregulation and cardiometabolic risk, we assessed relationships between these factors. We analyzed cross-sectional associations between arsenic, lead, mercury, cadmium, and cardiometabolic health markers in 178 randomly selected African-origin adults (52% female, 51% obese, mean age=43.0±6.4 years) from Ghana, South Africa, Seychelles, Jamaica, and USA. Metal levels were dichotomized to high or low at the median level of each metal. We analyzed associations between gut microbiome taxa, metal levels, clinical measures (BMI, fasting blood glucose, and blood pressure) and diagnoses (hypertension, obesity, and diabetes status). High vs. low lead and arsenic exposures had a significant effect on beta diversity (p <0.05). 71 taxa were associated with high lead levels: 30 with elevated BMI, 22 with T2DM, and 23 with elevated fasting blood glucose (p<0.05). 115 taxa were associated with high arsenic levels: 32 with elevated BMI, 33 with T2DM, and 26 with elevated blood glucose (p<0.05). Of the taxa associated with high lead and arsenic exposure and either elevated BMI or fasting blood glucose, porphyrin metabolism was the most enriched metabolic pathway. These data collectively provide the first findings in a human study that the gut microbiome may drive the association between lead and arsenic exposure and obesity and T2DM risk.

## INTRODUCTION

Type 2 diabetes mellitus (T2DM) and obesity are increasing worldwide health challenges associated with a large disease burden, comorbidities, and healthcare costs.(1-3) Globally, it is projected that 2.7 billion adults will be overweight or obese by 2025, and between 2025 and 2045, people with T2DM will increase by 212 million to 783 million.(1,3) By 2045, Northern Africa, currently highest in regional T2DM prevalence at 16.2%, is expected to increase to 136 million people, while sub-Saharan Africa is expected to have the highest T2DM prevalence increase of 129% to 55 million people.(1) Black Americans are disproportionately affected by obesity and T2DM, contributing to significant health disparities in the US.(4) While obesity is a main cause of T2DM, both obesity and T2DM also increase the risk of other highly prevalent cardiometabolic diseases (CMDs).(5) Successful strategies for management and treatment of obesity and T2DM require a more complete understanding of the risk factors that drive the complex heterogeneous etiopathology of these diseases.

Mounting evidence suggests environmental exposures, including toxic metals/metalloids (hereafter, “metals”) may contribute to CMD risk.(2-3,6-12) Metals (arsenic, cadmium, lead, and mercury) exposures come from food, water, and airborne sources.(2-3,8-13) For example, gold mining in Ghana has polluted river water with metals.(14) Arsenic, lead, and cadmium have been linked to increased risk of elevated fasting blood glucose (FBG), and arsenic has been linked to increased diabetes prevalence.(10-11) Although multiple mechanisms have been proposed, especially for arsenic, no specific mechanism has been well-defined.(2-3)

Gut microbiota, including their composition and microbially produced metabolites, are increasingly thought to be a significant player in the development and progression of obesity and T2DM.(15-22) The degree of dysbiosis appears to be associated with both obesity and T2DM disease severity.(15-21) In animal models, fecal microbiota transfer from individuals with T2DM or obesity can replicate the disease phenotype.(15,18-21) Overall, gut microbiota has been linked to metabolic disease development by multiple different mechanisms, including inflammation, gut barrier integrity, and microbial metabolites that act as signaling molecules.(22)

Recent evidence in animal models indicates that metals exposure may be linked to gut microbiome dysfunction with little data from human studies.(23) In the animal studies, metals have been suggested to drive dysbiosis through altered microbial composition subsequently impacting host physiologic processes and metabolic functions of the gut microbiome.(9,24-26) Lead, cadmium, and arsenic exposure have each been associated with decreased microbial diversity and specific differentially altered genera.(13,24-26) Metal exposures are also linked to altered metabolism of vitamins, bile acids, and other biomolecules and cofactors, where key steps in production of active biomolecules occur through microbial metabolism impacting host metabolism.(13,25-26) Understanding metals exposure impact on the human gut microbiome and critically, how this influences cardiometabolic health is needed.

To our knowledge, this study explores for the first time the association between toxic metals exposures, gut microbiome, and CMD in adults of African descent from five countries across the epidemiologic transition. We analyzed the difference between individuals with high versus low metals exposures across all sites as well as within sites. We hypothesized that high metals exposures and gut dysbiosis interact to promote metabolic dysregulation and increase CMD risk. These data reveal firsthand evidence of important associations between metals exposures and the gut microbiome and, in turn, associations with the prevalence of obesity and T2DM in critically impacted human populations.

## METHODS

### Study population

The main study population consists of an international cohort of 2,506 African-origin adults with ∼500 participants from each site: Ghana, South Africa, Seychelles, Jamaica, and USA. Participants are followed through the Modelling the Epidemiologic Transition Study (METS) (NIH R01-DK080763) and the currently funded METS-Microbiome study (NIH R01-DK111848). (27-28) METS-Microbiome explores the relationship between gut microbiota and T2DM and obesity risk.(27) The study collects urine samples, stool for gut microbiota sequencing, and clinical labs for measures of cardiometabolic health, which include T2DM risk (fasting insulin and glucose levels), metabolic health (lipids), and kidney function (urine creatinine).(27-28)

METS sites were selected as representative of the epidemiologic transition continuum, which is a model that captures the transition from predominantly infectious diseases to non-communicable diseases that accompanies increasing economic development.(27-29) The epidemiologic transition framework is defined using the United Nations Human Development Index (HDI) to study health outcomes across all sites with Ghana representing lower-middle-income, South Africa representing middle-income, Jamaica and Seychelles representing high-income, and the US representing very high-income.

Individuals were excluded from participating in the original METS study if they self-reported being persons with an infectious disease, including HIV, being pregnant, breastfeeding, or having any condition that prevented the individual from participating in normal physical activities.(27-28) Participants were further excluded from METS-Microbiome if they self-reported using antibiotics in the preceding 3 months. A description of both the METS and METS-Microbiome protocols for data collection, measurement, and laboratory procedures has been published.(27-28) Both METS and METS-Microbiome studies were individually approved by the Institutional Review Board of Loyola University Chicago, IL, US, which is the coordinating center.(27-28) For international sites, the protocols were approved by their respective institutions.(27-28) All study procedures were explained to participants in their native languages, and participants were provided written informed consent after being given the opportunity to ask any questions and compensated for their participation.

### Patient and public involvement

Patients and/or the public were not involved in the design, conduct, reporting, or dissemination plans of this research.

### Urinary metals and microbiome assessments

Urine samples were obtained in 2019 from 178 METS-Microbiome participants randomly selected and analyzed for metals levels (Table 1). Urine samples were tested to determine concentrations of arsenic, cadmium, lead, and mercury. To assess metals exposure impact on the microbiota in this subset, we accessed the 16S rRNA sequence data from 2019 and the measured clinical phenotypes to evaluate the most significant associations.

**Table 1.**
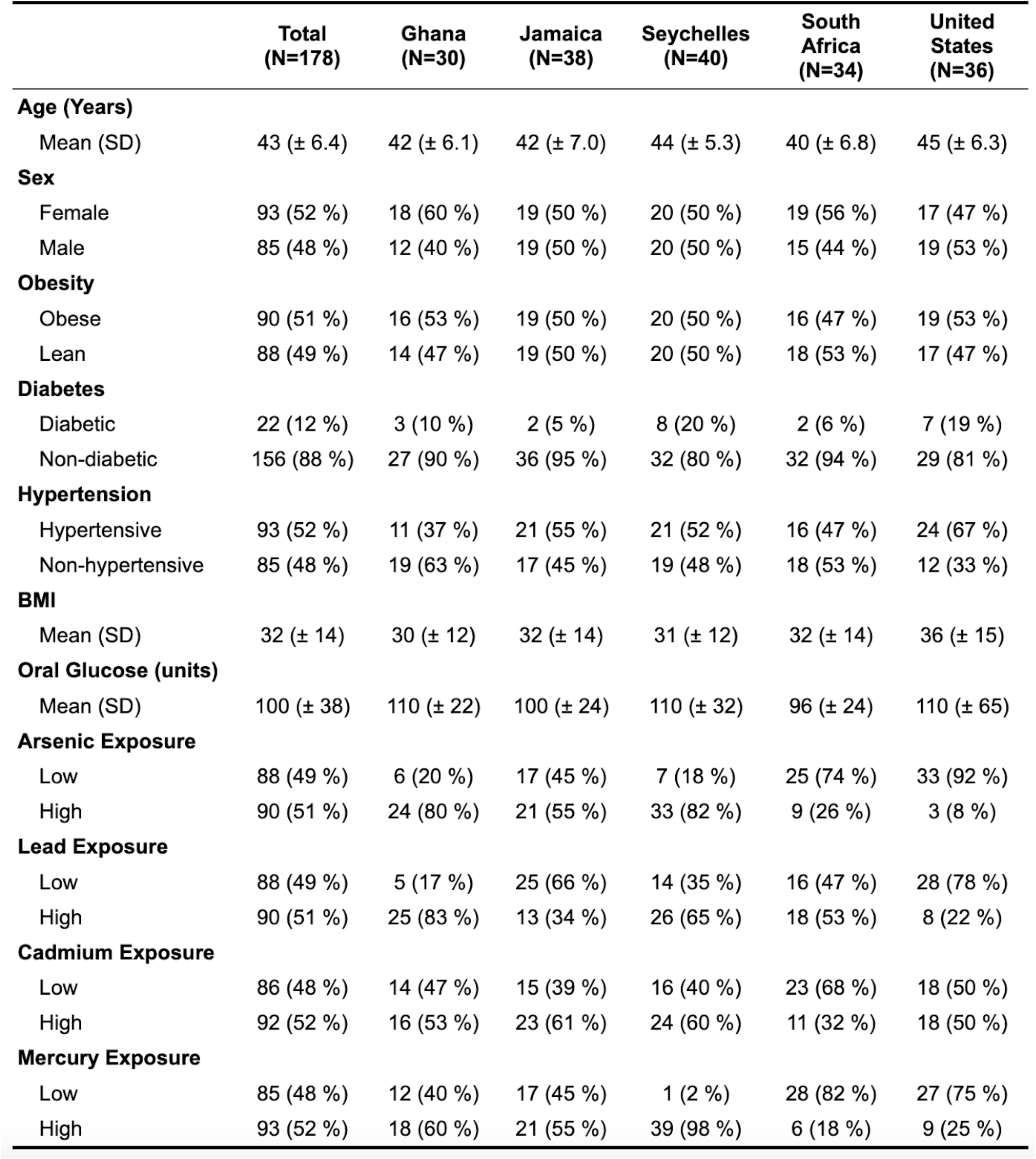
Characteristics of study participants for all sites. Aggregate data of metal exposure and cardiometabolic risk factors by site and across the dataset. Count and percentage of the population reported for categorical variables. Mean and standard deviation is reported for continuous variables.

### Anthropometry, sociodemographic, and biochemical measurements

METS visits were conducted at community-based research clinics within the respective communities, early in the morning following an overnight fast. Weight measurements were captured, BMI calculated, and obesity defined as BMI >30. FBG was measured; insulin, leptin, and adiponectin were measured in fasting plasma samples using radioimmunoassay kits (Linco Research, Inc., St. Charles, MO). T2DM was defined as FBG >125 mg/dL or by current treatment; except for Ghana, where not all participants fasted overnight, T2DM was defined as glucose ≥140 mg/dL or current treatment following American Diabetes Association guidelines for random glucose testing.(23-24) Blood pressure (BP) was measured in triplicate at two timepoints during each examination using an automatic digital monitor (model HEM-747Ic, Omron Healthcare, Bannockburn, IL USA). Spot urine samples were assayed for urinary albumin and creatinine levels.

### Urinary metals quantification

Urine samples were analyzed for arsenic, cadmium, lead, and mercury using inductively coupled plasma-mass spectrometry (ICP-MS). Urinary creatinine was measured using a method based on the Jaffe reaction for standardizing metal concentrations to control for kidney function and hydration status.(30) The core laboratory participates in the Quebec Multielement External Quality Assessment Scheme of the Institut National de Santé Publique du Québec, Canada for external accuracy assessment of biological samples. To assess the metals effect, each metal was stratified by median metals level per site for a within-site metals exposure label and by median metals level across all sites (the study population median) for a between-site metals exposure level.

### DNA extraction and amplicon sequencing

Fecal samples were sent to the Microbiome Core sequencing facility (University of California, San Diego, UCSD) and randomized for 16S rRNA gene processing to extract DNA with MagAttract Power Microbiome kit using blank controls and ZymoBIOMICS mock controls (Cat. No. D6300) in each extraction plate.(15,27) The V4 region of 16S rRNA gene was amplified from extracted DNA with 515F-806R region-specific primers according to the Earth Microbiome Project.(31) Purified amplicon libraries were sequenced on the Illumina eq platform to produce 150 bp forward and reverse reads (IGM Genomics Center at UCSD).(15,27)

### Bioinformatic analysis

We analyzed microbiota amplicon data using existing pipelines to identify taxonomic markers for all samples. In Qiita, generated raw sequence data was demultiplexed, quality filtered, and trimmed.(15,27,32) Amplicon Sequence Variants (ASVs) were defined using DeBlur, and taxonomy was assigned using a Naïve-Bayes classifier compared against a SILVA (version 138) reference database.(15,33) Microbiota samples were matched to study participants for whom clinical and urine metals metadata were available. The resulting ASV abundance count table, taxonomy data, and sample metadata were exported and merged into a phyloseq object in R (R Foundation for Statistical Computing, Vienna, Austria) for downstream analysis.(15) The phyloseq object was then used for quality control to remove: 1) ASVs with less than ten reads in the entire dataset and samples with fewer than 5000 reads; 2) ASVs that were unassigned at the phylum level; and 3) ASVs with fewer than 50 reads across all samples or were in less than 2% of samples. Quality control resulted in 16399 ASVs with 9264 genus-level taxa across 178 participants.

### Alpha and beta diversity analysis

Microbial alpha diversity was measured using the Shannon index via the microbiome library.(15) Beta diversity was calculated with pairwise Bray-Curtis dissimilarity, and significance was calculated with permutational multivariate analysis of variance (PERMANOVA) using phyloseq.(15) Univariate comparisons were performed in two-sample two-tailed t-tests when we could assume normality, and Wilcoxon Signed Rank tests when we could not. Benjamini-Hochberg (BH) adjusted p-values of less than 0.05 were considered statistically significant.

### Linear discriminant analysis

Linear discriminant analysis (LDA) effect size was performed on per-sample normalized relative abundances.(34) This algorithm estimates microbial taxa that contribute to observed differences by metals exposure.(34) We evaluated differences by coupling a univariate non-parametric test (Wilcoxon rank-sum, α = 0.05) with LDA scores (threshold for discriminative features > 3.0) to calculate effect size of identified differentially abundant taxa stratified by each individual categorical metal exposure metadata.

### Association analysis of microbial taxa

To identify multivariable associations within these data, four linear mixed effect models were used with individual taxa, urinary metal levels, and cardiometabolic risk profiles with MaAsLin2.(35) The ASV abundance table was transformed with trimmed mean of m values and run with a negative binomial model.(35) For each metal, a model measured individual associations {taxa ∼ metal + cardiometabolic variables + confounders}. A final model included all metal variables {taxa ∼ arsenic + lead + cadmium + mercury + cardiometabolic variables + confounders}. The cardiometabolic variables include obesity, T2DM, and hypertension diagnosis, with FBG levels, systolic and diastolic BP, and BMI. Each model controlled for site, sex, and age as confounders. From these four models, associations are calculated between taxonomic relative abundance at the genus level, metal exposures, and cardiometabolic variables. P-values were corrected for multiple comparisons using the BH correction, α=0.05.

### Predicted metabolic gene pathway analysis

Genera significantly associated with metals exposure were used to predict (q-value <0.05 to reduce false discovery rate) functional metabolic pathways. We utilized The Phylogenetic Investigation of Communities by Reconstruction of Unobserved Species 2 (PICRUSt2) v2.5.1.(36) Normalized ASV abundance table and weighted nearest-sequenced taxon index values per sample were used for predicting Enzyme Commission numbers and annotated using the MetaCyc database to identify enriched metabolic pathways.(36) The resulting enriched metabolic pathway abundances were visualized.^29^ P-values were adjusted for multiple comparisons using the BH correction, α=0.05.

## RESULTS

We analyzed data from 178 METS Microbiome study participants (median age = 43.0±6.4 years, 52% women) (Table 1). Median (IQR) BMI was lowest in Ghana, 20.82 kg/m^2^ (19.08, 38.36), and highest in the US, 30.12 kg/m^2^ (22.63, 49.26). 14.3% of the total study population had T2DM, with the highest (20%) in the US and Seychelles and the lowest (5%) in Jamaica. The highest lead levels were in Ghana, median (IQR) = 1.36 µg/g (1.11, 1.76) (83% had elevated levels), and lowest in US, 0.53 µg/g (0.38, 0.75) (22% had elevated levels). Arsenic levels were highest on average in Ghana; however, more participants had high arsenic exposure in Seychelles. In Ghana, median (IQR) arsenic level was 72.96 µg/g (39.5, 108.09) (80% had elevated levels) and in Seychelles, median (IQR) arsenic level was 61.71 µg/g (41.15, 97.0) (82% had elevated levels). Arsenic levels were lowest in the US at 7.99 µg/g (6.01, 12.66) (8% had elevated levels). Cadmium levels were highest in Jamaica, 0.508 µg/g (0.29, 0.90) (61% had elevated levels), and lowest in South Africa, 0.25 µg/g (0.16, 0.41) (32% had elevated levels). Mercury levels were highest in Seychelles, 1.98 µg/g (1.38, 2.54), (98% had elevated levels) and lowest in South Africa, 0.064 µg/g (0.007, 0.18), (18% had elevated levels) (Table 1).

### Alpha diversity

Gut bacterial diversity and richness measured by the Shannon alpha diversity index varied by country of origin, being higher in Ghana and South Africa, and lower in Seychelles, Jamaica, and the US (Fig. 1E). However, microbial diversity and richness between high versus low metal levels were only significant for high lead exposure in Seychelles and high cadmium exposure in the US (Fig. 1A-D). Overall, metals in our cohort had a minimal effect on alpha diversity.

**Figure 1.**
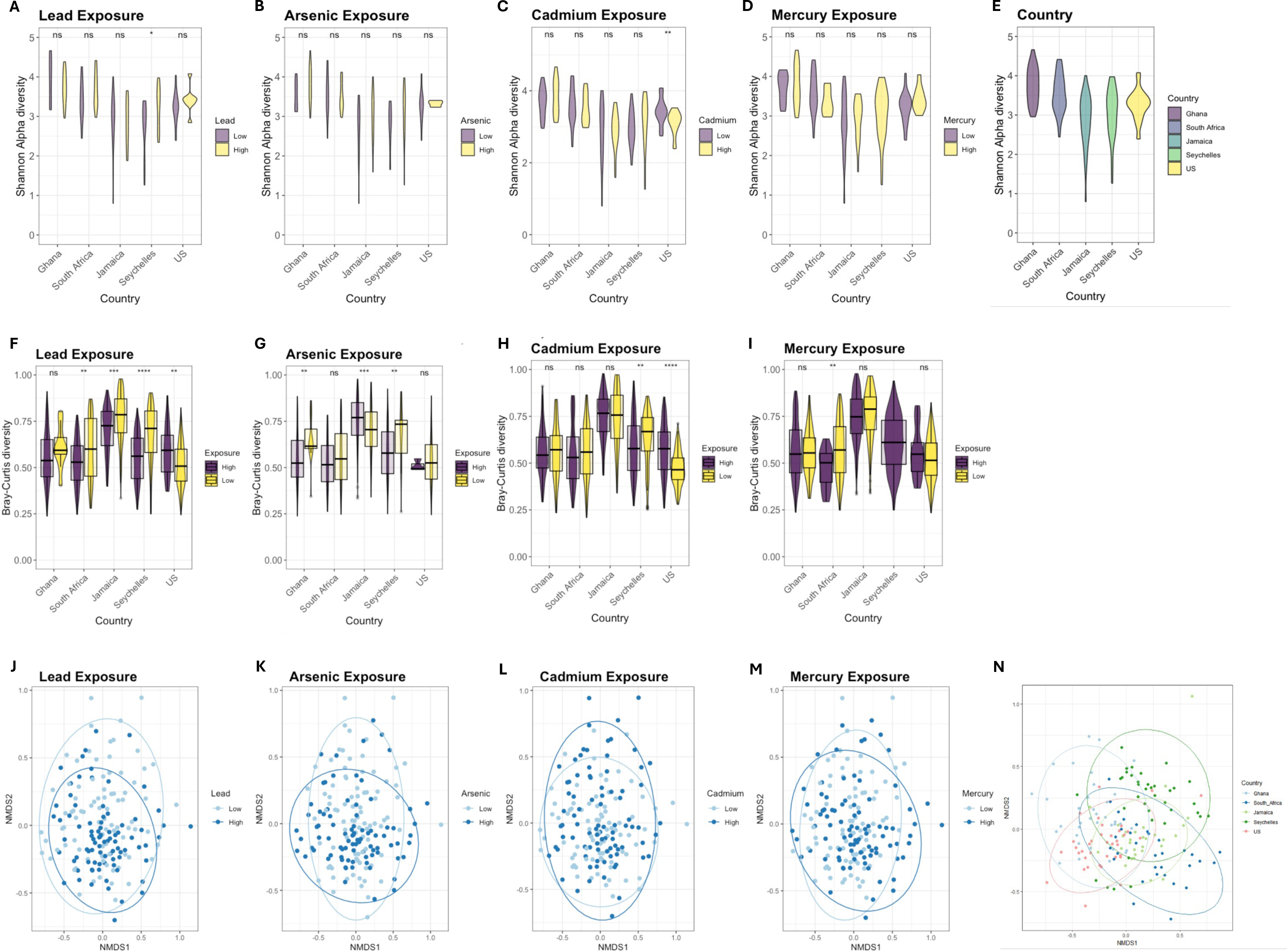
A-E) Shannon alpha diversity by metals exposure between countries of origin. Alpha diversity was significantly distinct by **A)** high lead exposure in Seychelles and **C)** high cadmium exposure in the US, but no significance existed by site with **B)** high arsenic exposure and **D)** high mercury exposure. **E) Alpha diversity across sites.** Alpha diversity is highest in Ghana and South Africa, and decreases in Jamaica, Seychelles, and USA. All associations are significant except between Jamaica and the US, Ghana and South Africa, and Jamaica and Seychelles. **F-I) Beta diversity by metals exposure between countries of origin. F)** high vs low lead exposure was significant for South Africa, Jamaica, Seychelles, and USA. **G)** High vs low arsenic exposure was significant for Ghana, Jamaica, and Seychelles. **H)** High cadmium exposure was significantly different from low exposure for Seychelles and USA. **I)** High vs low mercury exposure was significant for South Africa. Beta diversity was measured by the Bray Curtis dissimilarity metric. **J-N) Beta diversity across all countries**. Across all sites, **J)** high vs low lead exposure and **K)** high vs. low arsenic exposure were significantly different by PERMANOVA (p-value < 0.05) when controlling for country of origin**. L)** High vs. low cadmium exposure and **M)** mercury exposure were not significant by PERMANOVA (p-value < 0.05) when controlling for country of origin. **N) Country beta diversity across sites.** By PERMANOVA, there are significant differences (p-value <0.05) between Ghana and all other sites, South Africa and Seychelles, South Africa and USA, Seychelles and USA, Seychelles and Jamaica, and USA and all other sites. **** adjusted p < 0.0001, *** adjusted p<0.001, ** adjusted p<0.01, * adjusted p <0.05, ns p >0.05, paired Wilcoxon test.

### Beta diversity

Differences in bacterial composition or beta diversity demonstrated Ghana and the US were both significantly different from all other sites and Seychelles was significantly different from South Africa and Jamaica (Fig. 1N). Comparing high to low metal exposure groups, beta diversity was significant for lead and arsenic (Fig. 1J-M). By site, beta diversity significantly decreased in the high lead exposure group in South Africa, Jamaica, and Seychelles and increased in the US (Fig. 1F). In the high arsenic exposure group, beta diversity decreased in Ghana and Seychelles and increased in Jamaica (Fig. 1G). For the high cadmium group, beta diversity decreased in Seychelles and increased in the US (Fig. 1H). For the high mercury group, beta diversity decreased in South Africa (Fig. 1I).

### Linear discriminant analysis

With high lead exposure, *Clostridium*, *Subdoligranulum*, *Ruminococcus,* and *Peptostreptococcales* were the most differentially abundant taxa (Fig. 2A). With low lead exposure, no taxa were differentially abundant (Fig. 2A). *Firmicutes* and *Proteobacteria* were the primary phyla differentially abundant in high arsenic exposed microbial communities (Fig. 2B). The taxa overrepresented with high arsenic exposure include *Prevotella*, *Proteobacteria*, *Gammaproteobacteria*, *Enterobacterales*, *Christensenellaceae*, *Alloprevotella*, and *Clostridales* (Fig. 5B). With low arsenic exposure, differentially abundant taxa included *Anaerostipes*, *Erysiplatoclostridiaceae*, *Fusicatenbacter*, and *Ruminococcus* (Fig. 2B). Mercury-exposed microbial communities were only differentially characterized by *Anaerostipes* (Fig. 2C). High lead and both low and high arsenic drive differential regulation of associated taxa.

**Figure 2.**
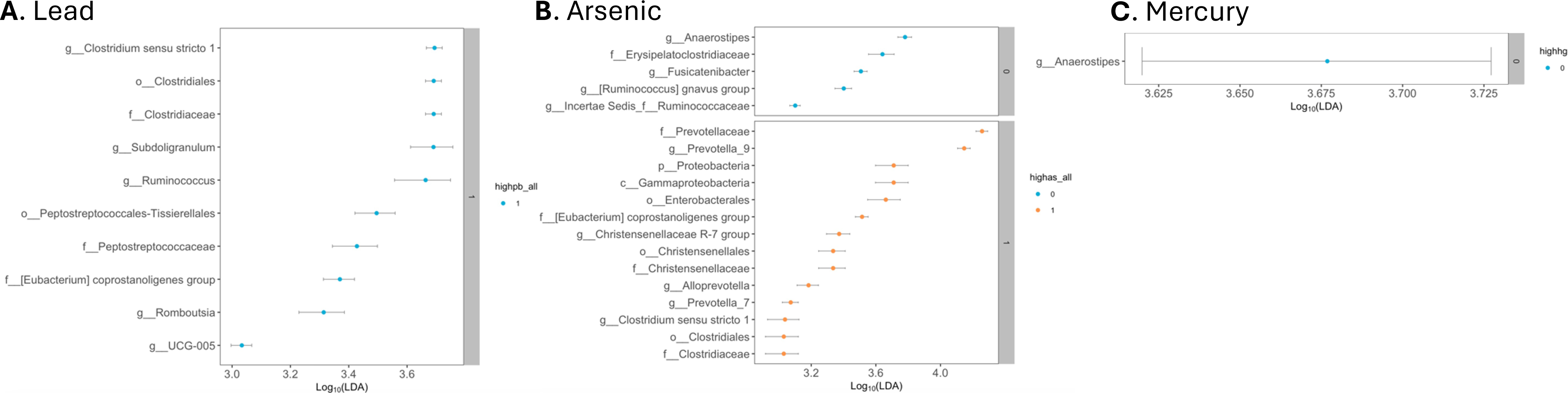
Differential taxa by metal exposure. **A)** High lead exposure characterized by several taxa. Low lead exposure not characterized by any taxa. Lead denoted as “highpb_all” with high lead categorized as “1”; low as “0”. **B)** High arsenic-exposed communities are unique compared with lower exposure. Arsenic is “highas_all” with high arsenic categorized as “1”; low as “0”. **C)** Low mercury-exposed microbial communities were only differentially characterized by one taxon. High mercury exposure not characterized by any taxa. Mercury is “highhg” with high mercury categorized as “1”; low “0”.

### Lead exposure

71 genera were significantly associated with high lead exposure: 30 strongly associated with BMI, 23 with FBG, and 22 with T2DM. Of these genera, 33 were significantly increased and 34 were significantly decreased in abundance with high lead exposure. High lead exposure increased phyla *Bacteroides* and decreased *Firmicutes*. At the genus level, high lead exposure was significantly associated with *Paraprevotella*, *Clostridium*, *Tyzzerella*, *Haemophilus*, and *Lachnospiracae* (Fig. 3A). High lead exposure was associated with changes in BMI (Fig. 3A) and increased obesity and T2DM incidence (Fig. 4A). For lead-associated taxa, BMI positively associated with *Clostridium*, *Lachnospiracae*, *Tyzzerella*, *Alloprevotella and Dialister*, and negatively associated with *Haemophilus*, *Neisseria*, *Rothia, Streptococcus*, and Family XIII AD3011 group. Lead-associated taxa positively associated with T2DM included *Clostridium*, *Haemophilus*, *Neisseria*, *Streptococcus, Family XIII AD3011 group*, and *Pediococcus* (Fig. 2D). Significant associations were found between high lead exposure, BMI, and hypertension for all lead-associated taxa, excluding *Haemophilus*, which associated only with high lead exposure and T2DM (Fig. 3A).

**Figure 3.**
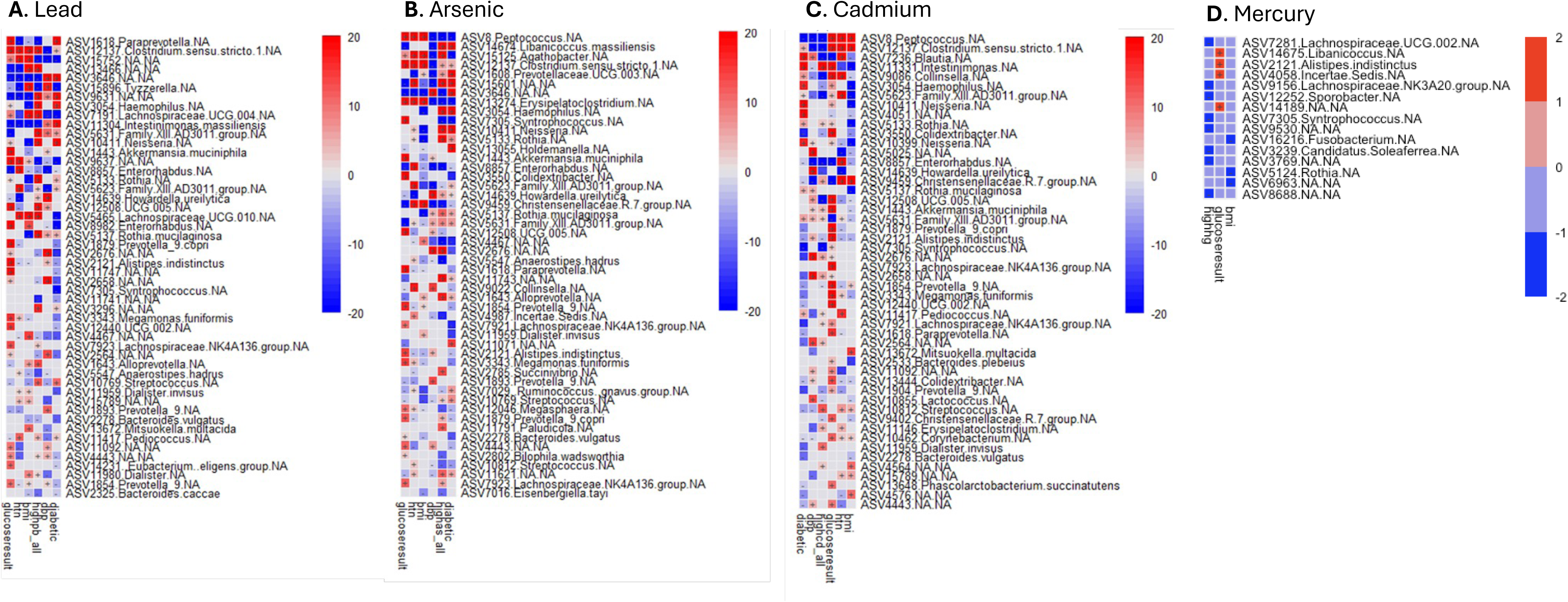
Differential taxa by individual metal exposure and cardiometabolic disease (CMD) factors. **A)** Lead and CMD factors, **B)** Arsenic and CMD factors, **C)** Cadmium and CMD factors, and **D)** Mercury and CMD factors. In each linear mixed model, taxa associated with metal exposure, diabetes diagnosis (diabetic), obesity diagnosis (obesity), hypertension diagnosis (htn), fasting glucose result (glucoseresult), BMI (bmi), systolic blood pressure (sbp), and diastolic blood pressure (dbp). Significance was calculated by -log(qval)*sign(coeff).

**Figure 4.**
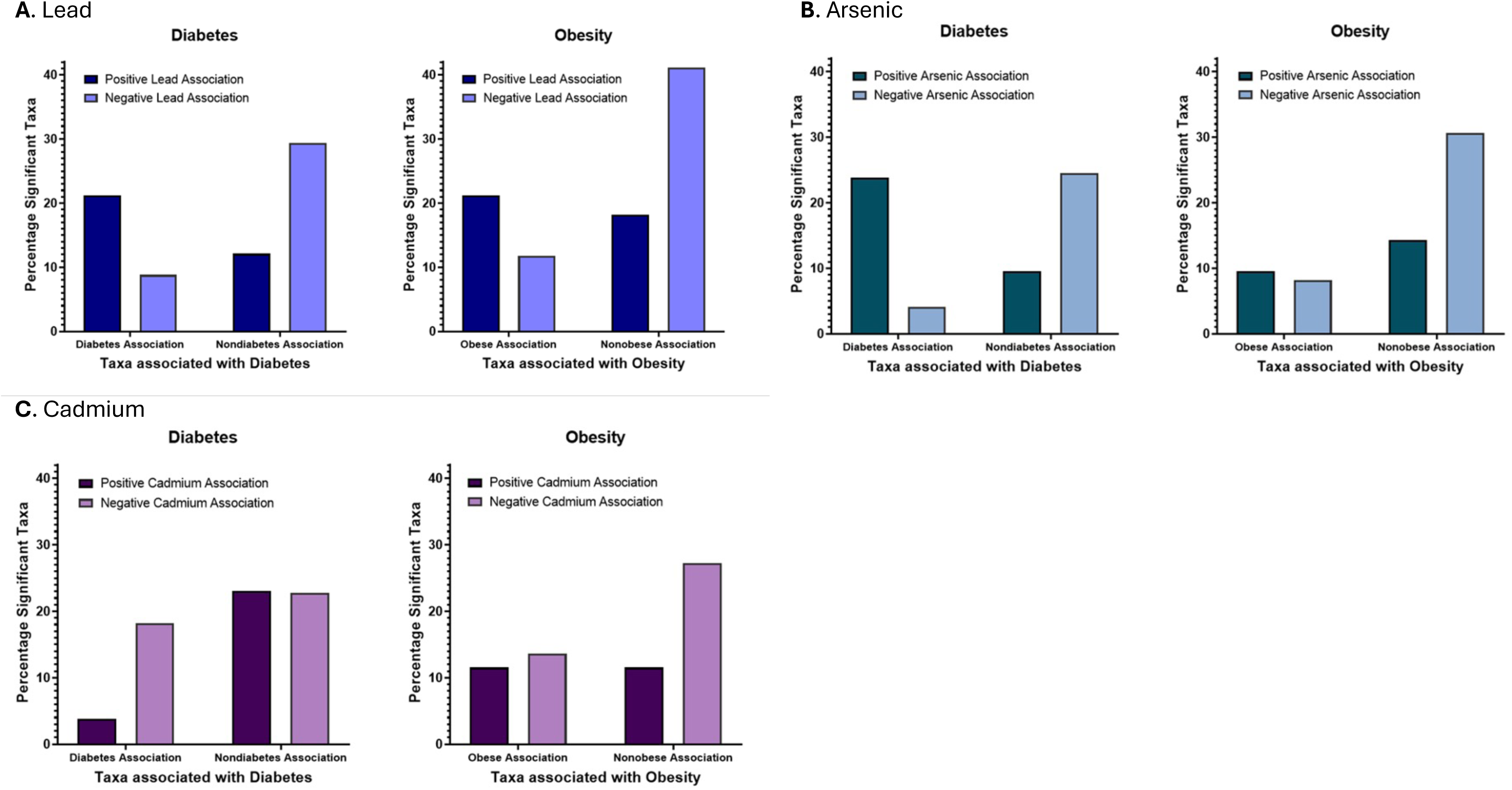
Percentage of significant taxa jointly associated with individual high metal exposure and obesity or diabetes. **A)** Taxa positively associated with lead exposure jointly associated with obesity and diabetes. More lead-associated taxa positively associated with diabetes (21% of taxa) than negatively associated (12%) (dark blue). More lead-associated taxa also positively associated with obesity (21%) than negatively associated (18%) (dark blue). Taxa that decreased abundance with high lead exposure were positively associated with a healthy phenotype and negatively associated with obesity (41%) and diabetes (29%) (light blue). **B)** More arsenic-associated taxa positively associated with diabetes (24%) than negatively (10%). 10% of taxa are positively associated with obesity and 14% negatively (dark green). Of the taxa that decreased with high arsenic exposure, more positively associated with a healthy phenotype and negatively associated with obesity (31%) and diabetes (24%) (light green). **C)** Of the taxa that increased with high cadmium; 4% positively associated with diabetes and 23% negatively associated (maroon). 12% of taxa positively associated with obesity and 11% negatively associated (maroon). Of the taxa that decreased with high cadmium; 23% negatively associated with diabetes and 27% negatively associated with obesity (pink).

**Figure 5.**
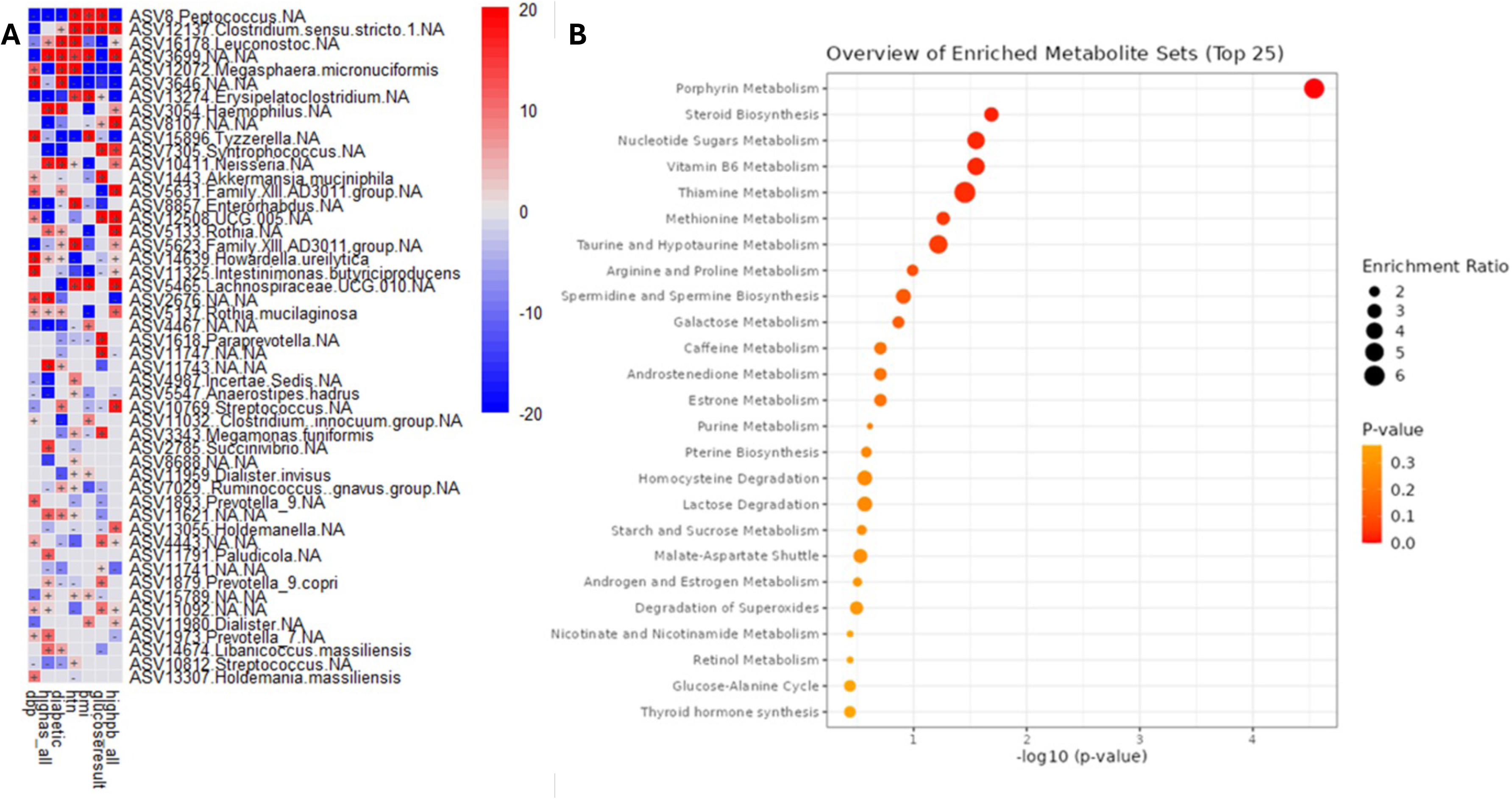
A) Top 50 taxa associated with all high metals exposures (lead, arsenic, cadmium, and mercury) and cardiometabolic factors, controlling for site, sex, and age. For the linear mixed model, taxa were associated with high lead exposure (highpb_all), high arsenic exposure (highas_all), high cadmium exposure (highcd_all), high mercury exposure (highhg), diabetes diagnosis (diabetic), obesity diagnosis (obesity), hypertension diagnosis (htn), fasting glucose result (glucoseresult), BMI (bmi), systolic blood pressure (sbp), and diastolic blood pressure (dbp), while controlling for sex, age, and site. Significance was calculated by -log(qval)*sign(coeff). **B) Top functional metabolic pathways.** Porphyrin metabolism is the most enriched metabolic pathway in the taxa associated with metal exposure. Steroid biosynthesis, nucleotide sugars metabolism, vitamin B6 metabolism, Thiamine metabolism, and methionine metabolism are all significantly enriched in this group (adjusted p value < 0.05).

21% of taxa were positively associated with the high lead group and an obesity diagnosis, compared to 18% associated with a lean phenotype. 21% of lead-associated taxa also associated with T2DM diagnosis compared to 12% with a non-diabetic phenotype (Fig. 4A). For taxa with a lower abundance in the high lead exposed group, 41% were associated with a non-obese phenotype (vs. 12% with obesity) and 29% were associated with non-diabetic phenotype (vs. 9% with T2DM) (Fig. 4A). Thus, high lead exposure resulted in an increased abundance in taxa associated with obesity and T2DM, and decreased abundance of taxa associated with nonobese and non-diabetic traits (Fig. 4A).

### Arsenic exposure

115 taxa were significantly associated with high arsenic exposure: 32 with BMI, 26 with FBG, and 33 with T2DM. Of these genera, 63 were significantly increased, and 49 were significantly decreased. High arsenic exposure increased abundance of taxa associated with T2DM (Fig. 3B and 4B). Genera that had the most significant positive association with high arsenic exposure were *Libanicoccus*, *Agathobacter*, *Haemophilus, Neisseria*, *Alloprevotella*, *Succinivibrio*, and *Rothia*. Arsenic was significantly associated with T2DM, and both high arsenic exposure, and T2DM were significantly associated with multiple taxa, but most significantly with Peptococcus, *Libanicoccus*, *Agathobacter*, *Clostridium*, *Prevotellaceae*, *Haemophilus*, *Syntrophococcus*, *Neisseria*, and *Rothia* (Fig. 3B). *Clostridium, Prevotellaceae,* and Alloprevotella were positively associated with both T2DM and BMI. *Neisseria*, *Haemophilus*, and *Rothia* were positively associated with T2DM and negatively associated with BMI. Finally, *Agathobacter* was shown to be positively associated with BMI and FBG, while negatively associated with T2DM (Fig. 3B).

### Cadmium exposure

48 genera were significantly associated with high cadmium exposure, with 26 positively associated and 22 negatively associated with exposure. Of the cadmium-associated genera, 14 were associated with BMI, 15 with FBG, and 15 with T2DM. The top genera that had the most positive association were *Intestinimonas*, and negative associations were *Peptococcus*, *Clostridium*, *Blautia*, *Collinsella*, *Enterorhabdus*, *Howardella*, and *Syntrophococus*.

High cadmium exposure decreased abundance of taxa associated with a nonobese or nondiabetic phenotype. Thus, high cadmium exposure was associated with obesity and T2DM (Fig. 4C). Of the top cadmium-associated genera, *Peptococcus* and *Clostridium* were positively associated with BMI while the rest were negatively correlated. *Clostridium*, *Blautia*, *Intestinimonas*, and *Collinsella* were positively correlated with T2DM, and *Peptococcus* and *Syntrophococcus* were negatively associated. High cadmium exposure and T2DM were significantly positively associated with *Intestinimonas, Rothia,* and *Family XIII AD3011 group;* the high cadmium group and T2DM were negatively associated with *Pepotcoccus, Pediococcus, Clolindextribactor*, *Enterorhabdus*, and *Syntrophococcus* (Fig. 3C).

### Mercury exposure

12 taxa were significantly associated with high mercury exposure, though none were jointly associated with any cardiometabolic variables (Fig. 3D).

### Comparing exposures and outcomes

To understand how each metal exposure interacts and associates with obesity and T2DM, we next ran a linear mixed model to identify microbial taxa associated with high metal exposures and clinical metabolic variables. *Peptococcus*, *Leuconostoc*, *Erysipelatoclostridium*, *Haemophilus*, *Tyzzerella*, and *Neisseria* significantly correlated with high lead exposure, high arsenic exposure, and T2DM. Additionally, high lead exposure alone positively correlated with BMI and had significant positive associations with *Clostridium* and *Megasphaera micronuciformis*. High arsenic exposure strongly correlated with 40% of the taxa significantly associated with T2DM. (Fig. 5A)

The results of this model demonstrated multiple genera significantly associated with high lead or high arsenic exposure are jointly associated with clinical measures of obesity and T2DM. High lead exposure was most strongly associated with abnormal FBG and elevated BMI, while high arsenic exposure was associated with T2DM and elevated diastolic BP. These results match with findings from previous mixed models for each individual metal (Fig. 3D-G), which showed high lead exposure associated most closely with obesity and high arsenic exposure associated with T2DM. (Fig. 5A)

### Metabolic pathways

From the microbes associated with metal exposure, increased BMI, and increased FBG, we found that porphyrin metabolism was the most highly enriched metabolic pathway and most significant by p-value (p < 2.1*10^-16^). Steroid biosynthesis, nucleotide sugars metabolism, vitamin B6 metabolism, thiamine metabolism, and methionine metabolism were all significantly enriched in this group (adjusted p< 0.05). (Fig. 5B)

## DISCUSSION

Lead and arsenic are known to be associated with obesity and T2DM, with high arsenic exposure associated with higher T2DM prevalence, as demonstrated in both human and animal studies.(2-3,6-12,23,37) Exposure to metals has previously been shown to have detrimental effects on the composition of the gut microbiota by both enhancing deleterious and suppressing beneficial taxa; however, most of these studies were done in mouse models. (9,13,24-26,38-40) A limited number of observational and retrospective studies have examined either arsenic or lead effects on the human gut microbiome, though none looked at cardiometabolic outcomes.(38,40) In mice, lead and arsenic have been shown to disturb gut microbiota by decreasing diversity.(25) As gut microbiota has been associated with metabolic risk, we assessed the linkage between toxic metals exposures and cardiometabolic outcomes in a diverse African-origin cohort. We found that two metals, lead and arsenic, are substantially associated with bacterial composition and impact T2DM and obesity. This study suggests that this increased risk is via changes to the gut microbiome possibly through the bacterial porphyrin pathway.

Regarding possible mechanisms for these associations, we found that arsenic depletes commensal bacteria in *Firmicutes*, including *Ruminococcus* and *Erysiplatoclostridiaceae*, both of which exhibit reduced abundance in the high arsenic exposure group and greater abundance in the low arsenic group. High-arsenic exposed microbiota show greater abundance of *Prevotella*, *Christensenella*, and *Clostridium*. In the high lead-exposed group, *Clostridium* and *Peptostreptococcales*, *Subdoligranulum*, and *Ruminococcus* had greater abundance. In sum, arsenic and lead exposure seem to differentially impact taxa, where these findings suggest that metals may increase the risk of obesity and T2DM by removing commensal taxa, possibly impacting metabolic protection provided through metabolites produced by the microbiota, and enriching taxa with pathogenic potential.^29^ The changes observed in the composition of the microbiota suggest that high metal exposure is associated with gut dysbiosis which contributes to increased risks of obesity and T2DM.

We found that the metabolism of the metal-associated microbial community is enriched in porphyrin metabolic pathways relative to taxa not associated with metals exposure. Porphyrin with an iron cofactor comprises heme and acts as an electron shuttle for the electron transport chain and as a molecule modulating redox signaling and stress, both critical in cellular respiration.(41-44) Notably, porphyrin metabolism plays a fundamental role in bacterial physiology as iron is a required cofactor for bacterial enzymes and proteins. Most bacteria have incomplete heme metabolic pathways and, therefore, take up available heme and iron via receptors and chelators.(43) Toxic metals exposure has been shown to affect heme synthesis through competition with iron by decreasing iron transport, reducing iron availability, and binding to proteins in place of iron.(44) Thus, exposure to toxic metals may impair heme synthesis and activity through a variety of mechanisms. With this impact on heme and iron availability in the gut lumen due to metals effects, bacteria with porphyrin metabolic pathways are upregulated, a process important to bacterial metabolism.(45) Interestingly, alterations in porphyrin metabolism and increased porphyrin metabolites have been associated with the development of insulin resistance and metabolic syndrome. (46-47) These changes additionally serve to drive gut dysbiosis and may be an additional factor in the downstream effects of gut dysbiosis on obesity and T2DM risk profiles.(48)

### Limitations

In this study, we have a single urinary estimate of toxic metal exposures, which may not reflect chronic exposures. For most metals, urinary measurements are well accepted for exposure evaluation.(49) While blood is generally utilized for evaluation of lead exposures, studies with blood and urine lead levels correlate with each other and urinary lead levels are considered a reasonable assessment for epidemiological studies.(50) Further, metal levels were binarized for a high vs. low comparison in our study, which will not capture any dose response to metals exposure or non-monotonic dose responses. For multiple analyses, we controlled for several potential confounders, but other measured and unmeasured variables may act as confounders for this data.

## CONCLUSION

Toxic metals exposures have been associated with obesity and T2DM previously, and, in animal models, these associations have been suggested to be partly driven by the gut microbiome. Our study of African-origin individuals has demonstrated for the first time, to our knowledge, that this could occur through changes in the gut microbiota. These metals effects may act by up-regulating taxa that are positively associated with obesity and T2DM and by down-regulating taxa that are negatively associated with obesity and T2DM. These data are critical as many countries are heavily impacted by deterioration in environmental quality. These results may inform strategies targeting the microbiome as a potential means of mitigating adverse metabolic effects of toxic metals, which may decrease the risk of obesity and T2DM, particularly in populations that are highly exposed to these metals. While reduction in exposures to detrimental substances is the cornerstone of environmental health interventions, interventions that can modulate the gut microbiome may help address the impact of unavoidable exposures. These observations provide new insight but warrant further validation and exploration.

## ETHICS STATEMENTS

### Ethics Approval

Both METS (IRB: LU200038) and METS-Microbiome (IRB: LU209537) studies were individually approved by the Institutional Review Board of Loyola University Chicago, IL, US, which also served as the coordinating center. For the international sites, the protocols were approved by the respective institutions and included the Committee on Human Research Publication and Ethics of Kwame Nkrumah University of Science and Technology, Kumasi, Ghana; the Research Ethics Committee of the University of Cape Town, South Africa; the Board for Ethics and Clinical Research of the University of Lausanne, Switzerland; the Health Research and Ethics Committee of the Ministry of Health of Seychelles; and the Ethics Committee of the University of the West Indies, Kingston, Jamaica. All study procedures were explained to participants in their native languages, and participants were provided written informed consent after being given the opportunity to ask any questions and compensated for their participation.

## Data Availability

The clinical and metadata are available under restricted access due to privacy regulations of our cohort. All data produced in the present study are available upon reasonable request to the authors. The SILVA 16 S rRNA database used for alignment is available at https://data.qiime2.org/2022.2/common/silva-138-99-515-806-nb-classifier.qza and the MetaCyc Databases are available at https://metacyc.org/.

## ACKNOWLEDGEMENTS

We would like to thank the staff and participants of the METS and METS-Microbiome study for their important contributions.

## Funding

This work was supported by the National Institutes of Health (R01 DK070853 and R01 DK111848 supporting AL and LRD; P30 ES027792 supporting RMS and MA; R01 ES028879 and R21 ES030884 supporting RMS; R01 DK104927 supporting BTL), the United States Department of Veterans Affairs (I01 BX00382 supporting BTL and I01 BX006108 supporting RMS), and the United States Department of Defense (TX220140 supporting RMS). Sequence data generated at the UC San Diego IGM Genomics Center with an Illumina NovaSeq 6000 was purchased with funding from a National Institutes of Health SIG grant under Award Number S10 OD026929 (J.A.G.).

## Competing Interests

No potential conflicts of interest relevant to this article were reported

## Author Contributorship Statement

J.A.J conducted the analyses and wrote the initial manuscript. C.C-K, A.L., K.B-A., T.F., P.B., E.V.L., D.R., and L.R.D. managed the METS-Microbiome study, sites, and sample collection. L.W. conducted metal quantification and contributed to drafting the methods. L.I. contributed to drafting the manuscript. J.A.G and G.E.M contributed to microbiome samples processing. C.C-K., J.A.G., G.E-M, A.L., P.B., E.V.L., D.R., T.K., M.A., L.R.D., R.M.S, Y.D., and B.T.L provided substantial contributions to the interpretation of the data and made critical revisions to the manuscript. All authors approved the final manuscript. Y.D. and B.T.L. contributed equally.

## Notes

### Competing Interest Statement

The authors have declared no competing interest.

### Author Declarations

Institutional Review Board of Loyola University Chicago, IL, US gave ethical approval for this work: METS (IRB: LU200038) and METS-Microbiome (IRB: LU209537). This was the central coordinating center for both projects. The Committee on Human Research Publication and Ethics of Kwame Nkrumah University of Science and Technology, Kumasi, Ghana gave ethical approval for this work. The Research Ethics Committee of the University of Cape Town, South Africa gave ethical approval for this work. The Board for Ethics and Clinical Research of the University of Lausanne, Switzerland gave ethical approval for this work. The Health Research and Ethics Committee of the Ministry of Health of Seychelles gave ethical approval for this work. The Ethics Committee of the University of the West Indies, Kingston, Jamaica gave ethical approval for this work. All study procedures were explained to participants in their native languages, and participants were provided written informed consent after being given the opportunity to ask any questions and compensated for their participation.

